# Quantitative transport mapping of multi-delay arterial spin labeling MRI detects early blood perfusion alteration in Alzheimer’s disease

**DOI:** 10.1101/2024.03.18.24304481

**Authors:** Yihao Guo, Liangdong Zhou, Yi Li, Gloria C. Chiang, Tao Liu, Huijuan Chen, Weiyuan Huang, Mony J. de Leon, Yi Wang, Feng Chen

**Affiliations:** Department of Radiology, Hainan General Hospital (Hainan Affiliated Hospital of Hainan Medical University), Haikou, China; Department of Radiology, Brain Health Imaging Institute (BHII), Weill Cornell Medicine, New York, New York, United States; Department of Radiology, Division of Neuroradiology, Weill Cornell Medicine, New York-Presbyterian Hospital, New York, New York, USA; Department of Neurology, Hainan General Hospital (Hainan Affiliated Hospital of Hainan Medical University), Haikou, China; Department of Radiology, MRI Research Institute (MRIRI), Weill Cornell Medicine, New York, New York, United States

**Author notes:** These authors contribute equally to this work and share the first authorship (Yihao Guo, Liangdong Zhou). **Corresponding authors:** Feng Chen, MD, PhD Department of Radiology, Hainan General Hospital (Hainan Affiliated Hospital of Hainan Medical University), No. 19, Xiuhua St, Xiuying Dic, Haikou, Hainan 570311, People’s Republic of China, Liangdong Zhou, PhD Department of Radiology, Brain Health Imaging Institute, Weill Cornell Medicine, 407 East 61St ST, New York NY 10066, United States.

**Keywords:** Cerebral blood flow (CBF), quantitative transport mapping (QTM), Alzheimer’s disease, early detection, cognitive function

## Abstract

**Background:** Quantitative transport mapping (QTM) of blood velocity, based on the transport equation has been demonstrated higher accuracy and sensitivity of perfusion quantification than the traditional Kety’s method-based blood flow (Kety flow). This study aimed to investigate the associations between QTM velocity and cognitive function in Alzheimer’s disease (AD) using multiple post-labeling delay arterial spin labeling (ASL) MRI.

**Methods:** A total of 128 subjects (21 normal controls (NC), 80 patients with mild cognitive impairment (MCI), and 27 AD) were recruited prospectively. All participants underwent MRI examination and neuropsychological evaluation. QTM velocity and traditional Kety flow maps were computed from multiple delay ASL. Regional quantitative perfusion measurements were performed and compared to study group differences. We tested the hypothesis that cognition declines with reduced cerebral blood flow with consideration of age and gender effects.

**Results:** In cortical gray matter (GM) and the hippocampus, QTM velocity and Kety flow showed decreased values in AD group compared to NC and MCI groups; QTM velocity, but not Kety flow, showed a significant difference between MCI and NC groups. QTM velocity and Kety flow showed values decreasing with age; QTM velocity, but not Kety flow, showed a significant gender difference between male and female. QTM velocity and Kety flow in the hippocampus were positively correlated with cognition, including global cognition, memory, executive function, and language function.

**Conclusion:** This study demonstrated an increased sensitivity of QTM velocity as compared with the traditional Kety flow. Specifically, we observed only in QTM velocity, reduced perfusion velocity in GM and the hippocampus in MCI compared with NC. Both QTM velocity and Kety flow demonstrated reduction in AD vs controls. Decreased QTM velocity and Kety flow in the hippocampus were correlated with cognitive measures. These findings suggest QTM velocity as an improved biomarker for early AD blood flow alterations.

## 1. Introduction

Alzheimer’s disease (AD) is the leading cause of dementia amongst elderly adults, that typically manifests prominent symptoms of progressive decline in memory and multiple other cognitive domains.^1^ The disease diminishes the quality of life for the patient, but also includes a heavy economic burden on society. Neurofibrillary tangles and amyloid-β neuritic plaques are the well-established pathological features of AD.^2,3^ However, AD pathogenesis is complex, involving multiple theories^4^, and includes multiple risk factors.^5^ Several studies have pointed out that vascular risk factors play an important role in the process of developing AD pathology.^6,7^ Previous studies have found that these vascular risk factors lead to vascular injury, resulting in cerebral perfusion alterations.^8^ Moreover, it has been shown that neuronal damage leads to reduced demand for oxygen and glucose, thus secondarily reduce the cerebral blood flow (CBF).^9,10^

Perfusion quantification using MRI is based on modeling a tracer transport through tissue captured in time-resolved imaging, such as dynamic contrast enhanced (DCE), dynamic susceptibility contrast (DSC), and arterial spin labeling (ASL). DCE and DSC methods use the injected gadolinium-based contrast agents (GBCA) as tracers. The ASL method, alternatively, uses radiofrequency labeled water as an endogenous tracer and is widely applied for perfusion quantification in clinical practice. Conventionally, these approaches for cerebral blood flow (CBF) quantification use Kety’s equation (Kety flow) by relating the temporal change in tracer concentration to an arterial input function (AIF) for each voxel.^11^ Since AIF at each voxel is not practically measurable, a single global AIF is assumed for blood flow to all brain regions, and is known to have errors and violate the local mass conservation principle.^12^ This commonly known AIF problem of conventional perfusion modeling has gained attention and encouraged development of new approaches using spatiotemporal information for perfusion quantification.^13^ To address this problem, we proposed to model changes in spatiotemporal tracer concentration according to the mass transport equation that utilizes spatial and temporal derivatives of the concentration without the selection of an AIF.^12^ Blood flow velocity can be calculated fully automatedly from fitting four dimensional (4D) dynamic tracer imaging data to the transport equation, which is termed as quantitative transport mapping (QTM).^12^ It has been demonstrated that 1) QTM velocity is more accurate than traditional Kety flow for blood flow quantification in in silico validation;^12,14^ 2) QTM velocity has a significant value in identifying breast cancer malignancy,^15^ nasopharyngeal cancer gene expressions^16^, lung shunt fraction^17^, and progressive liver disease stages.^14^

Given its promising diagnostic value in various diseases, we applied this technique to AD in this work for the first time to evaluate its ability of early detection. We tested two major hypotheses: 1) QTM velocity is superior to Kety flow in the separation of clinical AD spectrum groups; 2) QTM as compared with Kety’s method, offers better regional correlation with clinical cognitive performance measures.

## 2. Materials and Methods

### 2.1 Subjects

This study was approved by the Institutional Review Board (IRB) of our institute. All participants and/or their respective Legally Authorized Representative (when applicable) provided their written informed consent.

A total of 176 subjects aged 55 to 90 years old were recruited. All participants underwent neuropsychological tests and MRI examinations. Exclusions included 43 participants who were unable to complete neuropsychological tests and 5 participants who could not remain still in the MRI leaving 128 eligible subjects. A diagnosis of probable AD was made based on the criteria set by the National Institute on Aging and Alzheimer’s Association (NIA-AA)^18^ while a diagnosis of Mild Cognitive Impairment (MCI) was made according to Petersen.^19^ The definition of cognitively healthy control in additional to clinical interview was corroborated by Mini-Mental State Examination (MMSE) score > 27 and a Clinical Dementia Rating (CDR) score of 0.^20^

### 2.2 Neuropsychological Tests and Cognitive Outcomes

To assess cognitive status, 4 neuropsychological tests were administered.^21^

The MMSE is a 30-item screening tool used to summarize cognitive abilities including orientation, memory, attention, and language.^22^ We utilized the total score in our analysis.

The Trail Making Test (TMT) A and B require participants to draw a line connecting circles that contain numbers (A) or letters and numbers (B) in ascending order.^23^ The time needed to complete each test are indicators of processing speed and executive function.

In the Rey Auditory Verbal Learning Test (RAVLT), a list of 15 words is read 5 times. The participant is asked to recall the words after each presentation (immediate recall and learning). After a 20-minute delay, the participant is asked to recall the words again (delayed recall). We utilized the mean number of words recalled for the first 3 trials (immediate recall scores) as indicators of episodic memory and analyzed the total number of words recalled after the 20-minute delay (delayed recall score).^24^

The semantic verbal fluency test (VFT), the participant is asked to name as many animals as possible in 60 s. We utilized the total number of animals named as an indicator of semantic fluency.^25^

### 2.3 MRI data acquisition

All participants underwent MR examinations using a 3.0T MR scanner (Prisma, Siemens) with a 64-channel head/neck receiver coil. The imaging protocol included a three-dimensional (3D) magnetization-prepared rapid acquisition gradient-echo (MPRAGE T1w) sequence for anatomical imaging and a 3D pseudo-continuous ASL sequence with multiple post label delay (mPLD) for perfusion quantification.^26,27^ Scanning parameters were as follows: (1) MPRAGE: echo time (TE) = 2.26 ms; repetition time (TR) = 2300 ms; Inversion time = 900 ms; flip angle = 8°; slice thickness = 1 mm; field of view (FOV) = 256 × 256 mm^2^; voxel size = 1 × 1 × 1 mm^3^; (2) PCASL: TE = 37.78 ms; TR = 4200 ms; five PLDs = 500, 1000, 1500, 2000, 2500 ms; slice thickness = 3 mm; FOV = 240 × 240 mm^2^ ; voxel size = 2.5 × 2.5 × 3 mm^3^; Routine MR sequences (T2WI and T2-FLAIR) were also included to detect brain abnormalities.

### 2.4 MRI Data processing

#### 2.4.1 T1w based brain ROI parcellation

T1w MRI was regionally segmented using FreeSurfer (FS) version 7.1 ^28^ recon-all command for region of interest (ROI) parcellation. Individual ROIs defined by FS look-up-table (LUT) were combined bilaterally for the extraction of ROI values in Kety flow and QTM velocity maps. The Kety flow and QTM velocity were coregistered into FS T1w space before ROI value extraction. ROIs evaluated in this study include global cerebral cortex (GM), cerebral white matter (WM), deep gray matter (dGM), four cortical lobes (temporal (TL), frontal (FL), parietal (PL), occipital (OL)), and hippocampus (Hippo). To reduce the potential partial volume effect (PVE), all the ROIs used in this work were eroded 1 mm in FS space. We also have warped the Kety flow and QTM velocity into the Montreal Neurological Institute (MNI) space for the evaluation of group mean using volume-based Advanced Normalization Tools (ANTs) package. ^29^

#### 2.4.2 Kety flow mapping from multidelay ASL

Kety flow (ml/100g/min) maps were reconstructed from the mPLD PCASL data using the oxford_asl command in BASIL tools included in FSL.^30^ Specifically, the mPLD ASL data was first realigned using mcflirt in FSL with the M0 proton image as a reference.^31^ The realigned image was distortion corrected using the anterior-posterior and posterior-anterior encoding reference images with top-up correction implemented in FSL to reduce the effect of air-tissue boundary distortion in EPI-based sequence.^32^ The preprocessed mPLD ASL data was processed by subtracting the labeling image from the control image and then used the oxford_asl command for blood flow quantification with bolus time 1.5 s, PLDs = 0.5, 1, 1.5, 2, 2.5 s, T1 blood = 1.65 s, labeling efficiency = 0.85 and spatial smoothing regularization.^33,34^

#### 2.4.3 QTM velocity mapping from multidelay ASL

The quantitative transport mapping was modeled by the mass conservation equation of tracer^12,14,15^:

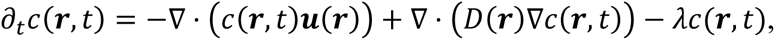

where *c*(***r****, t*) is the tracer concentration at location ***r*** and time *t*, ***u***(***r***) is the time-invariant tracer velocity, *D*(***r***) is the apparent diffusion coefficient, and *λ* is the signal decaying rate. For MR labelled endogenous water molecular in ASL data, the *λ* = 1/*T*1*b*, and *T*1*b* = 1.65s is the T1 time of blood. For perfusion estimation, *D*(***r***) could be considered negligible since diffusion effects are at much slower rate than blood perfusion. The reconstruction of perfusion velocity is then performed following the optimization below^12,14,15^:

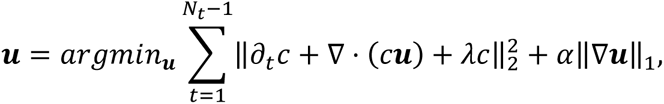

where α is the regularization parameter in the optimization to enforce a region-wise smooth solution. The reconstruction processing of ***u*** was performed using in-house code executed in MATLAB with realigned and top-upped multidelay ASL data. The magnitude of ***u*** was denoted as ||***u***||in L2 norm to represent the blood flow velocity in QTM.

### 2.5 Statistical Analysis

The statistical analyses were conducted using SPSS 20.0 and R Ver 4.3.2 in RStudio 2022. All significance tests were 2-sided with α = 0.05 as the significance threshold. Normality was assessed using the Shapiro-Wilk test for normality prior to testing group differences.^35^ Continuous variables were expressed as means ± standard deviations. Analysis of variance and χ^2^ tests were used to investigate group differences in demographic and cognitive variables. Kety flow and QTM velocity values of GM and hippocampus were compared among diagnostic groups using one-way analysis of covariance with age, sex, and years of education as covariates. Post-hoc multiple comparisons were performed to evaluate statistical differences between diagnostic groups. Correlation analyses were performed to investigate the relationships between regional Kety flow and QTM velocity across diagnoses. Finally, we assessed the effects of age and sex on blood perfusion and velocity using linear regression, and the association between cognitive score and perfusion measures using Pearson or Spearman’s correlation analysis. Note that all r reported are the correlation coefficient, and the p values reported are FDR adjusted for multiple comparison.^36^

## 3. Results

### 3.1 Clinical and demographic characteristics

Among 128 eligible subjects, there were 27 probable AD patients, 80 MCI patients and 21 NC. There was no significant difference in sex among the three groups (p = 0.213). Mean age was higher in the AD than the NC (p = 0.021) and MCI groups (p = 0.021), and years of education were higher in the NC than the MCI (p = 0.003) and AD groups (p = 0.002). Cognition scores including MMSE, immediate recall score, delayed recall score, TMT-A, TMT-B, and semantic fluency were significantly different among the three groups, consistent with the diagnoses (all p < 0.001). These results are summarized in Table 1.

### 3.2 Group mean of QTM velocity and Kety flow

Figure 1 presents the averaged maps of QTM velocity and Figure 2 shows the Kety flow for NC, MCI, and AD groups in the MNI space. The whole-brain patterns of Kety flow and QTM velocity across the diagnostic groups were visually different in AD from those in NC and MCI. Both of QTM velocity and Kety flow values across groups follow the order: NC > MCI > AD. The hippocampus region was highlighted by red arrow for perfusion comparison across groups.

**Figure 1.**
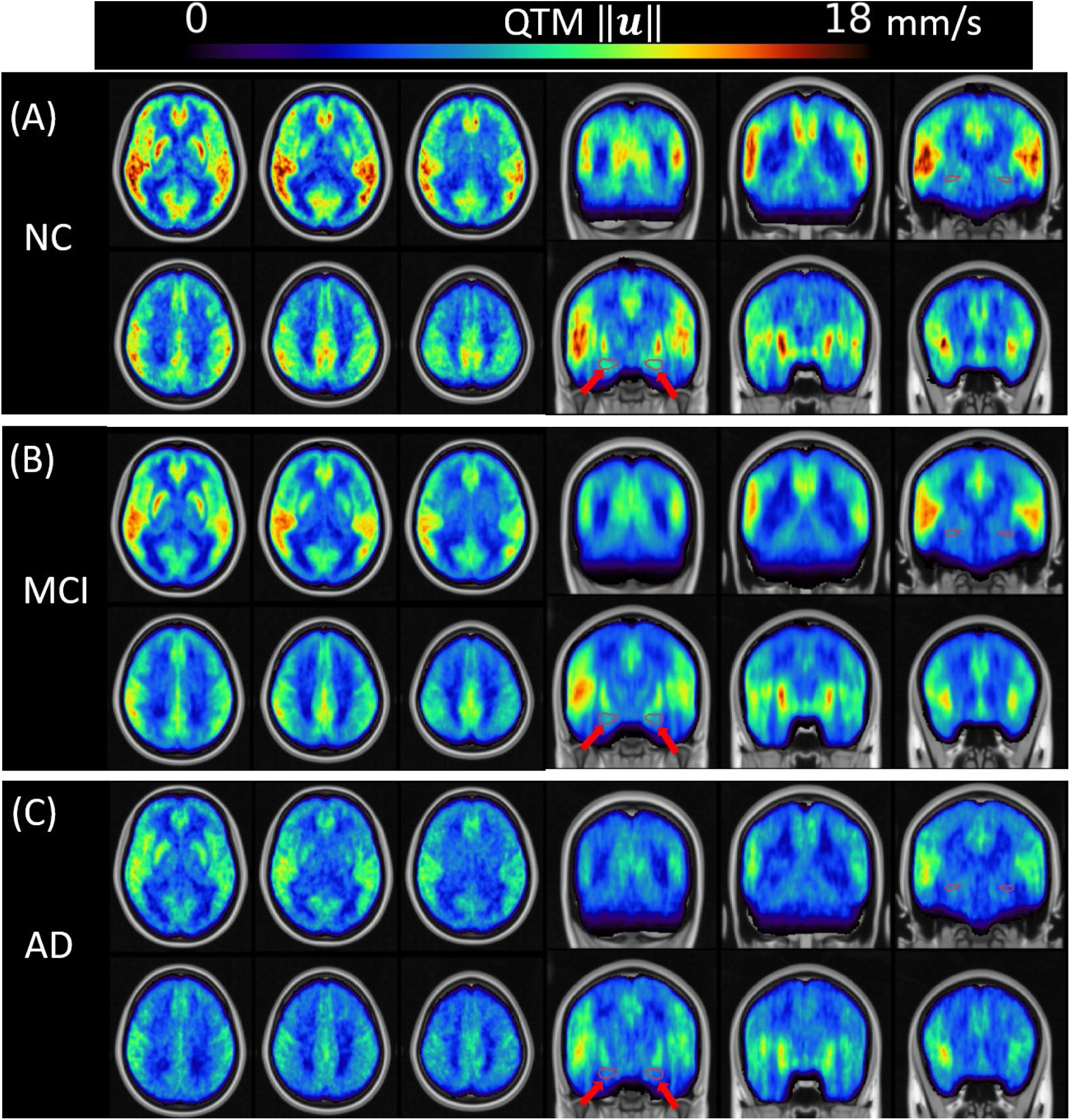
Group averaged QTM velocity in MNI template. (A), (B), and (C) are QTM velocity in NC, MCI, and AD, respectively. QTM velocity shows decreased pattern across NC, MCI and AD groups in axial and coronal views. Red arrows point to the hippocampus in coronal view to show the QTM velocity across groups. We see a drastically QTM velocity reduction in hippocampus from NC to MCI and AD.

**Figure 2.**
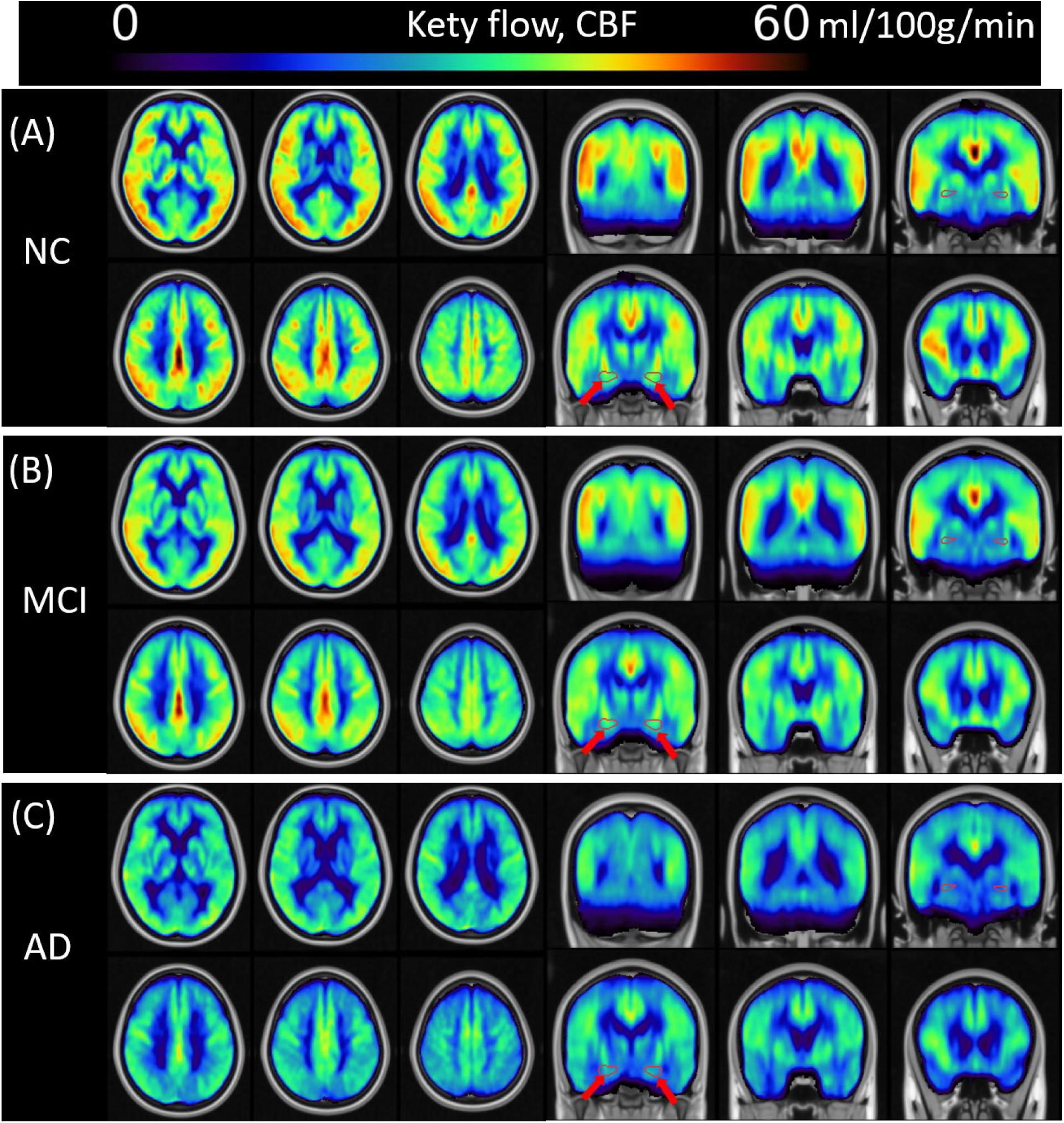
Group averaged Kety flow in MNI template. (A), (B), and (C) are Kety flow in NC, MCI, and AD, respectively. Kety flow velocity shows decreased pattern across NC, MCI and AD groups in axial and coronal views. Red arrows point to the hippocampus in coronal view to show the Kety flow across groups.

### 3.3 Group difference of regional QTM velocity and Kety flow

Figure 3 shows decreased QTM velocity values in MCI patients compared to NC in both GM (Figure 3(A), NC: 8.834 ± 1.758 mm/s, MCI: 8.006± 1.539 mm/s, p = 0.039) and Hippo (Figure 3(B), NC: 8.640 ± 2.253 mm/s, MCI: 7.360 ± 1.949 mm/s, p = 0.018). However, the Kety flow values were unable to distinguish MCI patients from NC (GM: p = 0.283, Hippo: p = 0.082). The results also revealed decreased QTM velocity value in AD patients compared to MCI and NC in GM (AD vs MCI: p = 0.027; AD vs NC: p = 0.001) and Hippo (AD vs MCI: p = 0.035; AD vs NC: p < 0.001), and decreased Kety flow value in GM (Figure 3(C), AD vs MCI: p < 0.001; AD vs NC: p < 0.001) and Hippo (Figure 3(D), AD vs MCI: p < 0.001; AD vs NC: p < 0.001).

**Figure 3.**
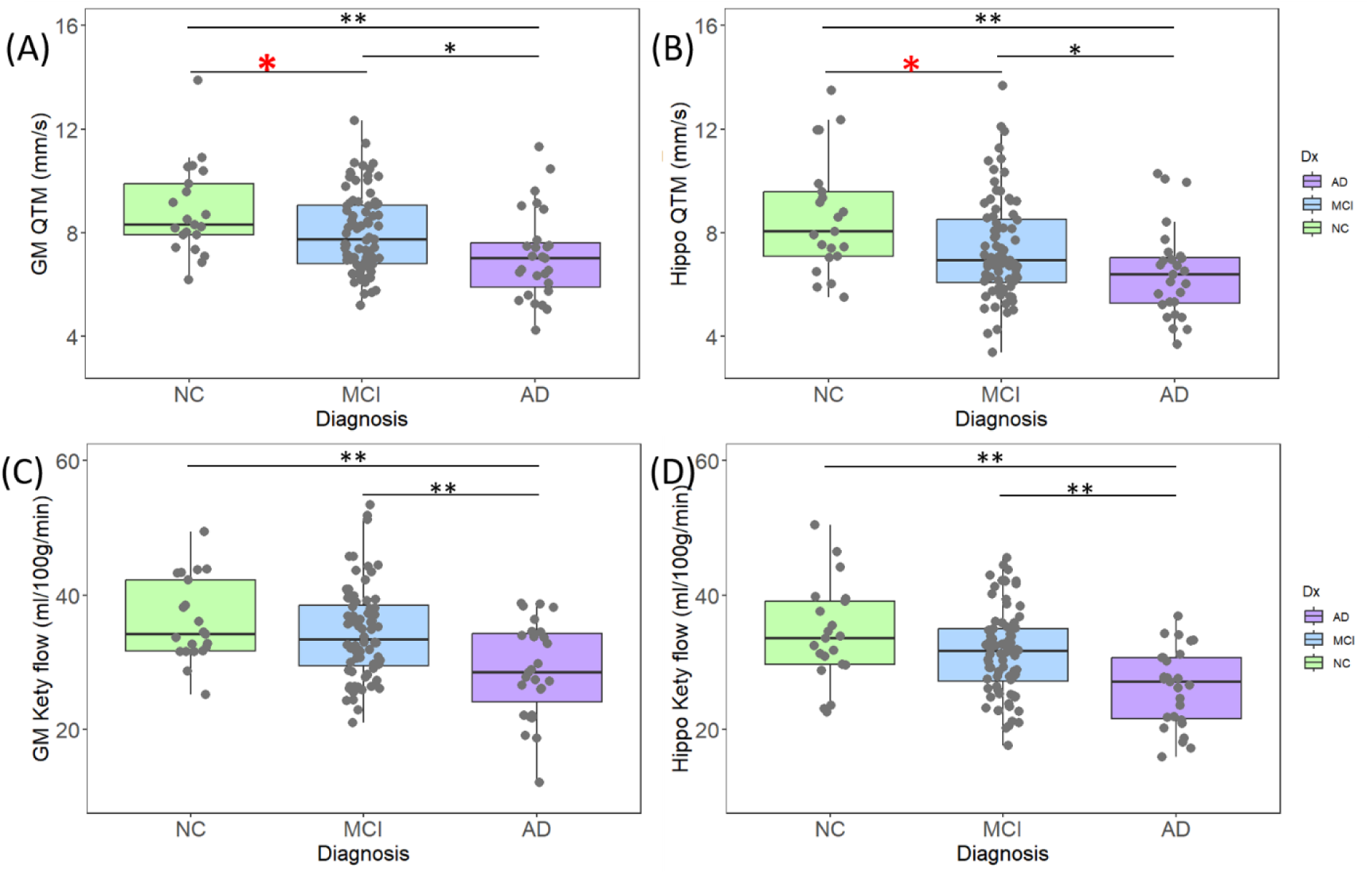
Pairwise group differences of regional QTM velocity and Kety flow. (A) and (B) are QTM velocity in GM and Hippo, respectively. (C) and (D) are Kety flow in GM and Hippo. QTM velocity shows group differences between NC, MCI and AD in both regions. Kety flow only differs between AD and MCI or NC, but not between MCI and NC. Asterisks indicates significance: * indicates p < 0.05, ** indicates p < 0.01. The red asterisks highlight the significantly QTM velocity reduce from NC to MCI, which was not applied to Kety flow.

### 3.4 Age and sex effects of QTM velocity and Kety flow

The effects of age and sex on QTM velocity and Kety flow are presented in Figure 4 in GM and dGM. Linear regression shows that QTM velocity in both GM (Figure 4A) and dGM (Figure 4B) significant reduced with age and male sex, with female having higher QTM velocity values than males (GM: age t = -3.133, p = 0.002, sex t = 2.459, p = 0.015; dGM: age t = -2.999, p = 0.003, sex t = 2.611, p = 0.010). In contrast, Kety flow in GM only shows a significant age effect, i.e., GM (Figure 4C) Kety flow decreases with age (t = -3.128, p = 0.002) but no sex effect (t = 1.442, p = 0.152), and the effect of sex is only significant in the dGM (Figure 4D) with higher Kety flow in females (t = 2.405, p = 0.018) but no age effect (t = -1.326, p = 0.187).

**Figure 4.**
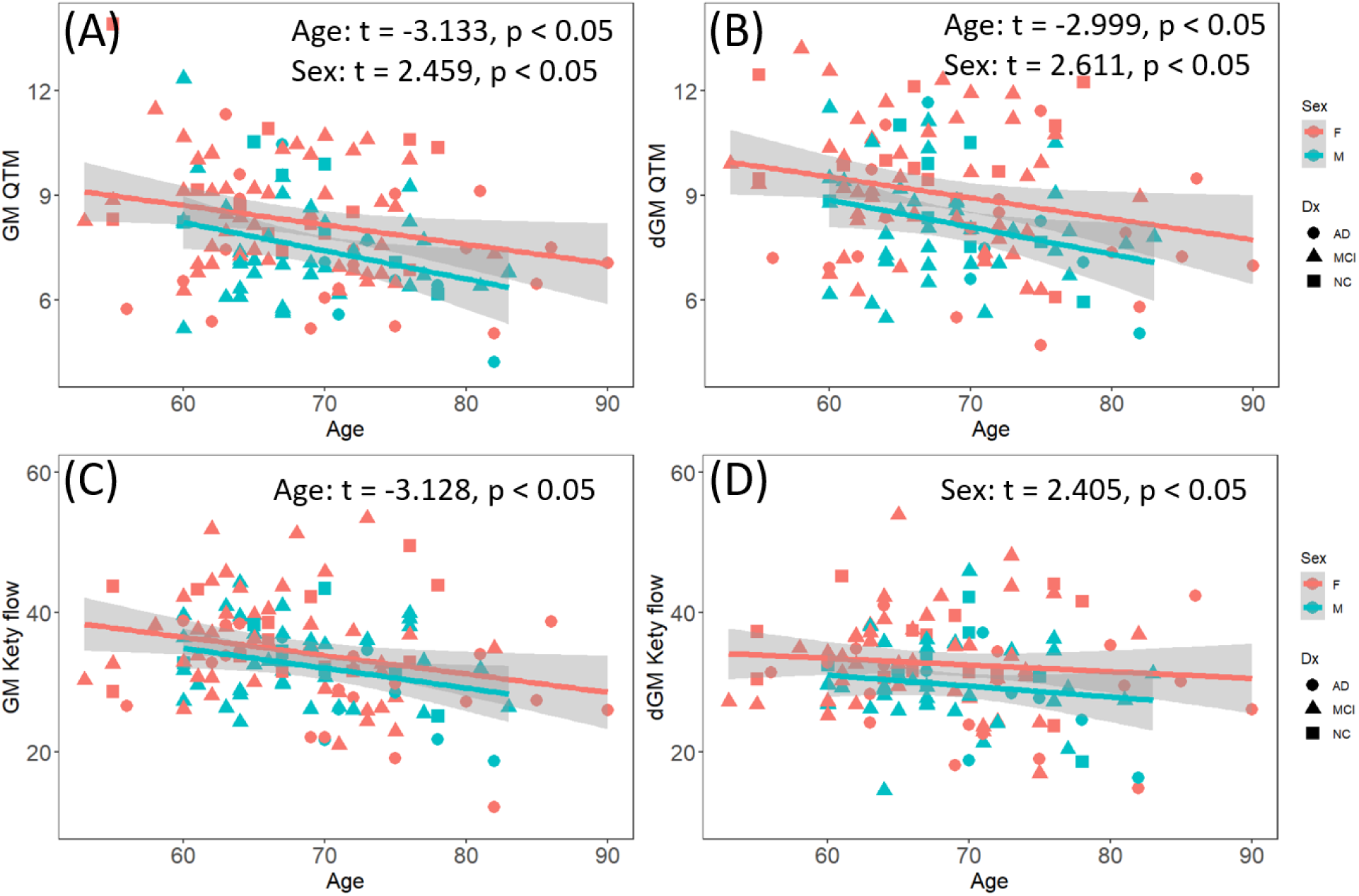
The age and sex effects of QTM velocity and Kety flow in cortical GM (GM) and deep GM (dGM). (A) GM QTM velocity with age; (B) dGM QTM velocity with age; (C) GM Kety flow with age; (D) dGM Kety flow with age. QTM velocity shows significant age and sex effects in both GM and dGM, while Kety flow only shows age effect in GM and sex effect in dGM.

### 3.5 Regional Kety flow and QTM velocity correlations

To investigate any inconsistencies between Kety flow and QTM velocity, we performed correlation analyses between regional Kety flow and QTM velocity across diagnostic groups. Results are shown in Figure 5. There were strong correlations between Kety flow and QTM velocity in the NC group in most regions, including GM, WM, FL, TL, OL, PL, Hippo, and dGM, and moderate correlations between Kety flow and QTM velocity in the MCI group. In the MCI group, there was a lower correlation between Kety flow and QTM velocity in almost all evaluated ROIs, demonstrating a mismatch between Kety flow and QTM velocity in these brain regions with disease progression. The correlations between QTM velocity and Kety flow in the AD group were generally lower than in the NC group except for PL, but similar or higher than in the MCI group.

**Figure 5.**
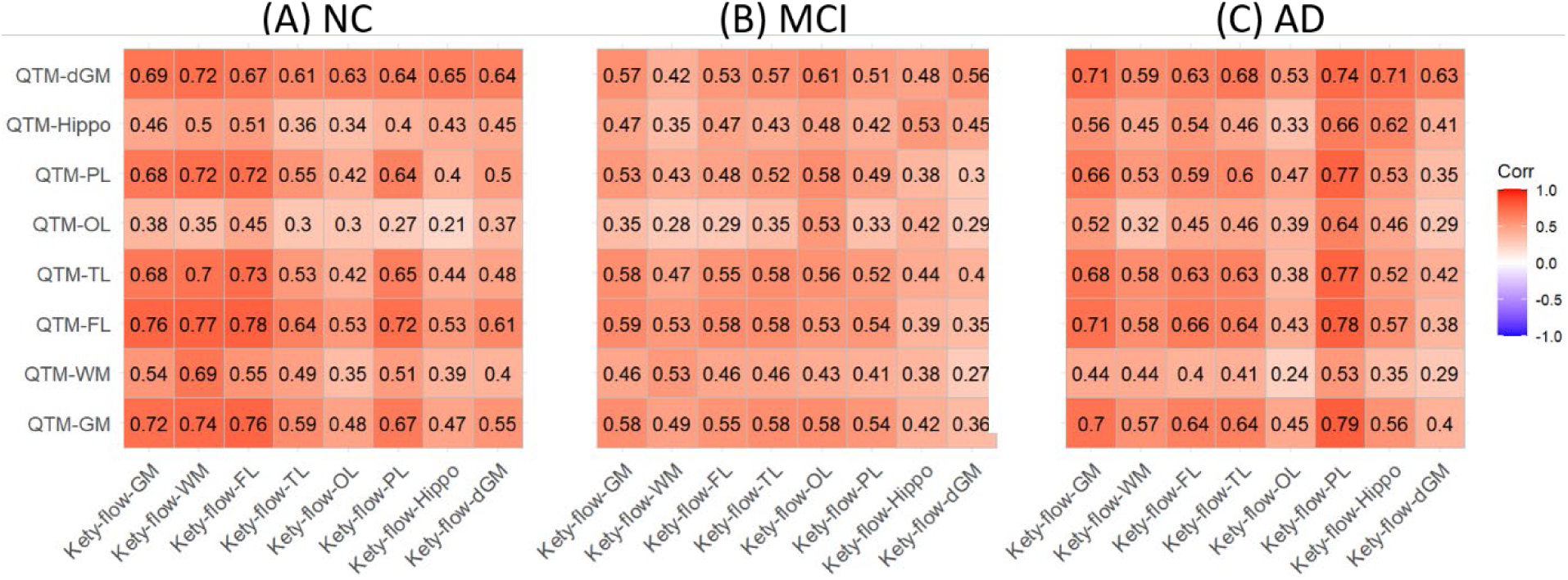
Correlation maps between QTM velocity and Kety flow in multiple regions, including GM, WM, FL, TL, OL, PL, Hippo, and DeepGM. The correlation is lower in MCI and AD groups compared to NC group. (A) correlation in NC group; (B) MCI group; (C) AD group.

### 3.6 Perfusion linked with cognitive function

Correlation analyses were performed to evaluate the relationship between cognitive abilities and perfusion measures in both GM and the hippocampus (see Table 2). Figures 6 (A)-(F) summarize QTM velocity with cognitive measures in the hippocampus, and Figure 6 (G)-(L) summarize Kety flow with cognitive measures. QTM velocity in the hippocampus was positively correlated with MMSE (Figure 6(A) r = 0.287, p < 0.01), immediate recall (Figure 6(B) r = 0.282, p < 0.01), delayed recall (Figure 6(C) r = 0.250, p < 0.01), and semantic fluency (Figure 6(F) r = 0.252, p < 0.01), and negatively correlated with TMT-B (Figure 6(E) r = -0.212, p < 0.01). Similar correlations were observed in GM for QTM velocity with cognitive measures and are summarized in Table 2.

**Figure 6.**
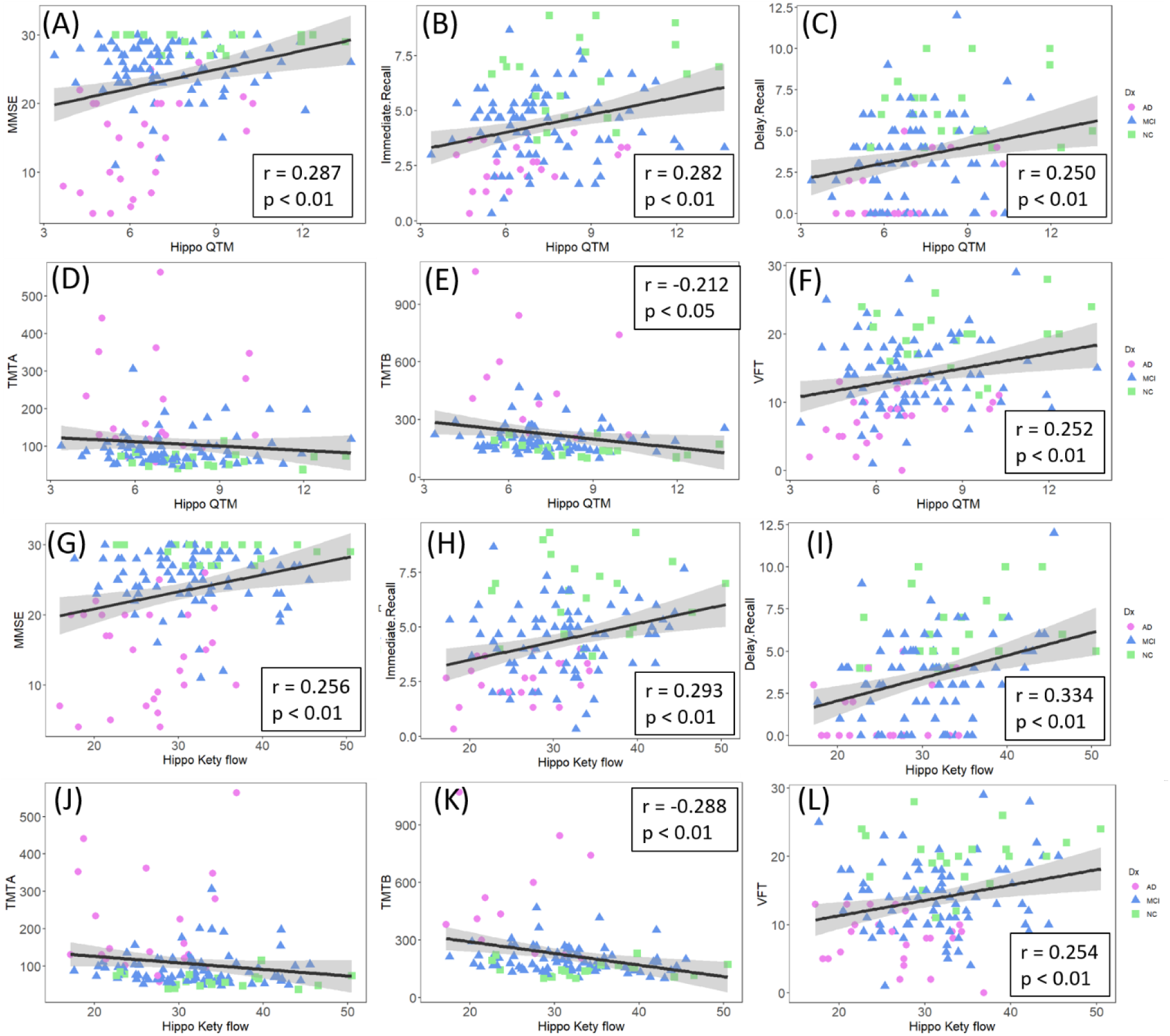
QTM velocity and Kety flow in hippocampus with cognitive measures. (A)-(F) are for QTM velocity; (G)-(L) are for Kety flow.

**Table 2.** Correlation of Kety flow and QTM velocity values with cognition in GM and hippocampus.

|  | MMSE |  | Immediate recall |  | Delay recall |  | TMT-A |  | TMT-B |  | Semantic fluency |  |
| --- | --- | --- | --- | --- | --- | --- | --- | --- | --- | --- | --- | --- |
|  | r | p | r | p | r | p | r | p | r | p | r | p |
| Relationship between CBF and cognition |  |  |  |  |  |  |  |  |  |  |  |  |
| GM | 0.29 | <0.01 | 0.27 | <0.01 | 0.28 | <0.01 | -0.22 | <0.05 | -0.35 | <0.001 | 0.26 | <0.01 |
| Hippo | 0.26 | <0.01 | 0.29 | <0.01 | 0.33 | <0.01 | NS | NS | -0.29 | <0.01 | 0.25 | <0.01 |
| Relationship between QTM and cognition |  |  |  |  |  |  |  |  |  |  |  |  |
| GM | 0.27 | <0.01 | 0.26 | <0.01 | 0.21 | <0.05 | NS | NS | -0.19 | <0.05 | 0.23 | <0.01 |
| Hippo | 0.29 | <0.01 | 0.28 | <0.01 | 0.25 | <0.01 | NS | NS | -0.21 | <0.05 | 0.25 | <0.01 |

Kety flow in the hippocampus was positively correlated with MMSE (r = 0.256, p < 0.01), immediate recall (r = 0.293, p < 0.01), delayed recall (r = 0.334, p < 0.01), and semantic fluency (r = 0.254, p < 0.01), and negatively correlated with TMT-B (r = -0.288, p < 0.05), as shown in Figure 6 (G)-(L). Kety flow in GM shows similar correlations and are summarized in Table 2.

## 4. Discussion

Our results demonstrate significant differences in QTM velocity between NC and MCI groups in the GM and hippocampus, while these differences are not observed with Kety flow, suggesting QTM velocity could constitute an early biomarker for AD. QTM velocity in GM and dGM is sensitive enough to detect perfusion changes regardless of age and sex. Additionally, our results show that both QTM velocity and Kety flow in the GM and hippocampus are significantly associated with overall cognition (MMSE), immediate recall score, delayed recall score, TMT-B and VFT. Cerebral perfusion, an indication of blood supply to tissue, is a potential early biomarker of AD and its alteration may appear earlier in AD than other hallmark pathological changes, such as beta-amyloid (Aβ) deposition, hyperphosphorylated tau accumulation, and widespread brain atrophy.^37,38^

### 4.1 Kety flow and QTM velocity for quantification of blood perfusion

Kety flow has been widely used to investigate perfusion changes in patients with MCI and AD. Considering patients with developed AD dementia, hypoperfusion was present in most of the brain areas, including GM, OL, FL, PL, TL, amygdala and hippocampus.^39–42^ We observed decreased Kety flow in GM and hippocampus for patients with AD than NC, which was consistent with previous studies. For comparisons between NC and MCI subjects, a meta-analysis study demonstrated that no obvious changes in Kety flow in the global, white matter, and GM were identified in MCI subjects.^43^ However, some previous studies demonstrated decreased Kety flow of MCI compared with NC in PL, OL, FL and TL ^41,42,44^. Our data show no significant difference in Kety flow between NC and MCI groups in both GM and dGM. These results demonstrate that Kety flow is not sensitive enough for detection of perfusion change in early stage of AD.

Previous works have shown that QTM velocity is able to distinguish benign from malignant breast lesions,^15^ and separate high grade nonalcoholic fatty liver disease (NAFLD) from low grade NAFLD using dynamic contrast enhanced (DCE) MRI, showing its high sensitivity to subtle physiological change at the early stage of diseases.^14^ In this work, QTM velocity calculated from mPLD ASL data is applied to detect cerebral blood flow change in AD. Our results found significant differences in QTM velocity between NC and MCI groups in both GM and hippocampus, demonstrating that QTM velocity could to be an early biomarker of AD.

### 4.2 Technical issues in the quantification of Kety flow and QTM velocity

Technically, Kety flow using a kinetic modeling with a global AIF as an input, and a lump of empirical parameters to evaluate blood flow in a voxelwise manner.^11,45^ The problems with Kety flow include its violation of local mass conservation due to its use of global AIF and its use of only temporal information by ignoring the spatial transport tracer between neighboring voxels.^13^ These simplifications of Kety’s model result in the loss of sensitivity of local and spatial change of blood flow, especially for patients with subtle pathophysiological changes like early stage AD.

In contrast, QTM model tries to solve both issues in Kety’s model by implementing biophysical mass transport model with dynamic data. First, QTM doesn’t require an AIF to quantify blood velocity by considering the spatiotempoal derivative of the 4D signal in the formula, which enables us to evaluate both spatial and temporal change in the dynamic data. Second, QTM doesn’t have assumed parameters that rely on empirical testing, which makes it suitable for all subjects including healthy and patients.

### 4.3 Perfusion measures associated with age and sex

Our results are consistent with previous studies in that Kety flow decreases with age in GM but not in dGM.^46^ QTM velocity decreases with age in both the GM and dGM, demonstrating a higher sensitivity for detecting perfusion changes than Kety flow. Kety flow values in the dGM are higher in females than males, while Kety flow in GM are relatively similar. A previous study shows that females exhibit significantly higher Kety flow values when compared to males.^47^ These contradicting results may be due to sample variation. The previous study includes subjects with aged 20 to 80, while our study includes older subjects with aged 55 to 90.^47^ These results indicates that perfusion is linked to age and sex, and that QTM velocity is more sensitive than Kety flow in detection of age- and sex-related alteration in perfusion. To eliminate these effects when comparing diagnostic groups, age and sex served as covariates.

### 4.4 Correlations between QTM velocity and Kety flow

We observed strong correlations (r > 0.55) between QTM velocity and Kety flow in the NC group across many evaluated ROIs. In the MCI group, this association decreased to moderate (r ∼= 0.45) in the same ROIs. As the disease progresses, the correlations between QTM velocity and Kety flow change across NC, MCI and AD. The potential reasons for this correlation change may be attributed to two main factors. First, the global AIF in Kety flow might be sufficient in NC subjects since their blood perfusion is high and the local estimated Kety flow is not affected much due to systematic estimation error. However, in MCI subjects with pathophysiological change in vasculature and perfusion, the disadvantage of global AIF starts to play a role and reduces the sensitivity and accuracy of Kety flow, due to alterations in blood flow pathway or quantity. On the other hand, QTM does not rely on AIF to estimate the blood perfusion and thus offers high sensitivity in estimating blood velocity and its subtle change. Second, the decreased perfusion in MCI patients compared with NC could be due to spatial changes in vasculature structure and blood perfusion routes. Kety flow fits the ASL data in Kety’s model with a voxelwise manner that only considers the temporal relationship between data frames, while QTM model based on biophysical principles, utilizes spatiotemporal information of dynamic data to explore the transport of tracer in blood across voxels. QTM model utilizes the same data as Kety’s model more efficiently, and may be the second reason why QTM is the more sensitive measure. From the correlation maps, we also observe that some brain regions are more affected than others during the disease development in AD, which needs further investigation of the blood route supply alteration in AD spectrum.

### 4.5 Decreased Kety flow and QTM velocity linked with cognition

The hippocampus is considered a major player in memory. Hippocampal atrophy is an established imaging biomarker in AD and constitutes neurodegeneration in the A/T/N framework.^48,49^ However, blood flow decline occurs much earlier than the appearance of brain atrophy.^50^ In the hippocampus, we found decreased Kety flow and QTM velocity in the AD group compared to the MCI and the NC groups. Moreover, QTM velocity showed a significant difference between the MCI and NC groups, demonstrating hippocampal perfusion changes at the early stage of AD, and was not seen using conventional Kety flow. Our results also showed that Kety flow and QTM velocity in the hippocampus correlated with cognition, including global cognition, memory, executive function, and language. These findings confirmed that blood flow coupled with blood flow velocity in the hippocampus can be useful diagnostic markers of AD.

Glymphatic function has been proposed to explain the brain clearance deficits in AD. Blood flow may play a key role in the general glymphatic flow, whose deficits has been shown to be the cause of Aβ accumulation. ^51–54^ QTM velocity from the labeled freely diffusible water signal in ASL may reflect the total fluid transport in the brain, including blood flow in vascular space and fluid flow in perivascular and interstitial spaces.^55^ The content of glymphatic flow information in QTM velocity derived from ASL would require further investigation.

### 4.6 Limitation of this study

We recognize several limitations in this study. First, the relatively small sample size in NC and AD groups. A large-scale prospective study, containing participants from subjective cognitive decline is needed to further explore the underlying mechanisms of blood flow velocity changes. Second, the ground truth of blood flow and blood flow velocity of the brain is unknow, although we had performed numerical simulations in kidney and liver to confirm the accuracy of QTM, a 3D printed phantom with known ground truth will be ideal to validate Kety’s method and QTM for brain perfusion quantification in the future. Third, PET imaging data is not available, thus the association between QTM velocity, brain clearance, and Aβ deposition is not yet investigated. Finally, the nature of cross-sectional study limits us to study different subjects at different disease stages. Our near goal is to run longitudinal study using QTM velocity to further study its underlying mechanism for early detection in AD. Future research will include PET data and explore the association between QTM velocity and brain Aβ deposition to design longitudinal studies.

## 5. Conclusions

This study demonstrated a reduced QTM velocity in GM and the hippocampus in MCI patients compared with NC, suggesting QTM velocity is a potential early biomarker for AD. Decreased Kety flow and QTM velocity in the hippocampus correlated with cognitive decline. These findings contribute to an improved understanding of perfusion change and cognitive decline in AD.

## Data Availability

All data produced in the present study are available upon reasonable request to the authors.

## 7. Acknowledgement

This work was supported in part by the Hainan Province Science and Technology Special Fund (ZDYF2024SHFZ058), the National Natural Science Foundation of China (82271977), the Hainan Academician Innovation Platform Fund, and the Hainan Province Clinical Medical Center, and the NIH of United States R01EB034755, R01AG080011, R01AG057848, R01AG068398, R01AG080011, RF1AG057570, R56AG058913, R01AG057848.

We would like to thank Emily B. Tanzi at Brain Health Imaging Institute (BHII) in Well Cornell Medicine for her efforts in the language proofreading.

